# Closed-loop Cortical Network Stimulation as treatment for refractory epilepsy originating from the primary motor cortex

**DOI:** 10.1101/2023.11.11.23298410

**Authors:** D. van Blooijs, S. Blok, E.J. Aarnoutse, N.E.C. van Klink, G.J.M. Huiskamp, M.D. Bourez-Swart, T.A. Gebbink, P. van Eijsden, S.M.A. van der Salm, N.F. Ramsey, F.S.S. Leijten

**Author notes:** Corresponding author: D. van Blooijs, UMC Utrecht Brain Center, Department of neurology and neurosurgery, University Medical Center Utrecht, PO Box 85500, 3584 GA Utrecht, The Netherlands. Disclosures: Medtronic (Minneapolis, Minnesota, United States of America) provided all components of the implant (subdural leads, extension leads, neurostimulator) and devices to set sensing and stimulation parameters (Sense Programmer, Clinician Programmer and antenna) free of charge, and provided technical support. Medtronic did not fund this study, the researchers or the patients in any other way.

## Abstract

**Background:** In epilepsy patients, cortical electrical stimulation is therapeutically applied in the seizure onset zone (SOZ) to reduce seizures. However, in patients with epilepsy arising from the primary motor cortex (M1), stimulation can result in undesired muscle contractions or loss of motor control. We postulate that seizure frequency reduction can also be obtained by cortical network stimulation in a site outside M1 with a connection to the SOZ in M1.

**Methods:** Patients with electroclinical seizures suspected to arise from M1 were selected. SOZ was delineated during chronic intracranial EEG monitoring. Using Single Pulse Electrical Stimulation, the underlying effective corticocortical network was determined and a site for stimulation was selected that was connected to the SOZ. One subdural strip was implanted on top of the SOZ, and one on the stimulus location. A subcutaneous neurostimulator (Activa^®^ PC+S, Medtronic), capable of recording and closed-loop stimulation, was connected to both strips. Seizure data was collected for three to five months and used to optimize a seizure detection algorithm. After this, closed-loop cortical network stimulation was applied during seven to nine months.

**Results:** In five subjects (two females, mean age 34 years, range: 21-51 years), a neurostimulation system was implanted. One subject was seizure free for 17 months post-implantation without applying any electrical stimulation. Two subjects were responders with a mean seizure frequency reduction of 73%. In two subjects, seizure frequency was reduced by on average 35%.

**Discussion:** In this clinical trial with five subjects suffering from refractory epilepsy arising in M1, seizure frequency was reduced with electrical stimulation in all subjects. This is a proof of concept showing that closed-loop cortical network stimulation can reduce seizure frequency as equal to direct SOZ stimulation in non-primary motor epilepsy.

## Introduction

Over the last decades, neurostimulation has become a treatment option that is regularly used in refractory epilepsy patients. With deep brain stimulation and vagal nerve stimulation, large networks in the brain are modulated resulting in seizure frequency reductions of 50% in around 50% of the patients ^1,2^. When there is a clear focal region responsible for epileptic seizures, more targeted, focal, cortical neurostimulation can be applied with better effects on seizure frequency reduction than large network stimulation ^1^. In a few case studies ^3^, cortical open-loop stimulation is applied to the epileptogenic region resulting in seizure frequency reductions of around 80-90%. With open-loop stimulation, neurostimulation is applied according to a pre-programmed pattern (e.g. 1 minute on, 5 minutes off) regardless of underlying brain activity.

With closed-loop stimulation, neurostimulation is applied when epileptic activity is detected. In a large trial applying closed-loop cortical neurostimulation, a responder rate of 73% and a mean seizure frequency reduction of 75% was observed ^4^. In patients with epilepsy arising from the primary sensorimotor cortex, stimulation in the seizure onset zone (SOZ) may lead to side-effects like twitches or sensations ^5^. Several of these cortical stimulation studies ^6,7^ mention that more research is needed regarding the stimulation site that is most effective for neurostimulation therapy, and that this site might not necessarily be the SOZ. We postulate that, instead of stimulating in the eloquent SOZ, stimulation in a directly connected, healthy area may be an effective alternative treatment strategy.

Recent studies ^8–11^ have shown that neurostimulation was more effective when the stimulation site had more connections with other regions and that the underlying network could be a predictor in effectiveness of stimulation therapy. Furthermore, we previously demonstrated^12^ that single pulse electrical stimulation in a connected region modulates interictal epileptic activity in the epileptogenic area, and suggested that this might be a good indicator for long-term neurostimulation and can be used to induce seizure reduction in areas unsuited for direct cortical stimulation, most notably the primary motor cortex. In this study, we investigate whether closed loop cortical network stimulation in healthy tissue connected to the SOZ in the primary sensorimotor cortex can reduce seizure frequency and improve quality of life.

## Methods

### Patients

In this prospective study, we included patients who were suspected of focal epilepsy arising from the primary sensorimotor cortex around the central sulcus. Patients had to be at least 16 years of age; with a seizure frequency of at least two seizures per day, and at least three anti-seizure medications tried.

We delineated the SOZ in detail by means of intracranial subdural EEG in order to be certain that it was indeed located within the primary sensorimotor cortex and was not eligible for surgery because of the risk of unacceptable functional deficits post-surgery. Candidates in whom this criterium was not met, underwent epilepsy surgery and were then excluded from the neuromodulation trial.

The REC2Stim (Rational Extra-eloquent Closed-loop Cortical Stimulation) study was approved by the ethics committee at the University Medical Center Utrecht and the national Dutch Health and Youth Care Inspectorate, in accordance with the Declaration of Helsinki (2013). This study was registered with clinicaltrials.gov (NCT04158531).

### Informed consent procedure

People were aware of, and conditionally assented to, alternative neuromodulation treatment in case the clinical invasive monitoring would reveal a non-resectable focus in the primary sensorimotor cortex. They were given full information on the REC2Stim neuromodulation trial, including its experimental part. Patients signed an intention to informed consent before undergoing invasive epilepsy monitoring, which technically counted towards study participation. Final informed consent was obtained only after clinical delineation of the SOZ in eloquent cortex. This approach was adopted because confronting the patient with the trial at the end of a clinical invasive epilepsy monitoring period would leave insufficient time for considerations and questions.

### Invasive epilepsy monitoring

Patients underwent clinical invasive epilepsy monitoring with subdural electrocorticography (ECoG) for 4-7 days. During implantation surgery, a trepanation was performed and electrode grids were placed subdurally over the pericentral area suspected of generating seizure activity. This area was determined with pre-surgical evaluation, including seizure semiology, MRI and video-EEG.

During this invasive epilepsy monitoring period, we visually analyzed spontaneously occurring seizures to delineate the SOZ, and applied electrical stimulation mapping to delineate motor and sensory functions. We applied Single Pulse Electrical Stimulation (SPES; ten monophasic, bipolar stimuli of 0.2 Hz, 4-8 mA, 1 ms) to each neighboring electrode pair. In our hospital, SPES is used as part of the clinical evaluation to localize the epileptogenic region ^13^. We reconstructed the underlying effective network ^14^ from the corticocortical evoked potentials (CCEPs) to SPES stimuli. In this network, we determined electrodes outside the eloquent region with connections towards the SOZ, and determined whether SPES stimuli in these extra-eloquent electrodes resulted in transient suppressive effects in the ongoing ECoG inside the SOZ. Electrode sites connected to the epileptogenic region and modulating activity in the SOZ on SPES stimulation were potential candidates for therapeutic stimulation after completion of invasive epilepsy monitoring. Details of this procedure are provided in the Supplementary Appendix and Supplementary figure 1.

### Selection of electrodes for seizure detection

Consensus between the responsible neurologist (FL) and the clinical neurophysiology team specified the SOZ electrodes based on visual inspection of the seizure data. We analyzed their interictal and ictal power spectrum. The electrodes that showed the largest difference between interictal and ictal power spectra were selected as the sensing site for seizure detection. Details of the electrode selection for seizure detection are provided in the Supplementary Appendix and Supplementary figure 2.

### Selection of electrodes for therapeutic stimulation

Based on their connection to the SOZ and neuromodulatory effects during SPES (see Supplementary Appendix and Supplementary figure 1), we selected three potential candidate sites in each patient for stimulation trials with various frequencies and current intensities during two days prior to grid explantation and implantation of the neurostimulator. We analyzed power spectra pre- and post-stimulation and determined the most effective of the three locations for therapeutic neurostimulation. Details of the stimulation protocol and of the electrode selection for therapeutic stimulation are provided in the Supplementary Appendix, Supplementary figure 3, and Supplementary figure 4.

### Implantation and description of the neurostimulation device

For this study, a neurostimulator (Implantable Pulse Generator, Activa PC+S^®^, Medtronic) with sensing capabilities was used. The Activa^®^ PC+S is an investigational device provided by Medtronic for use in clinical research studies. All components of this device, including electrode leads, were designated for investigational use. Details of the features of the neurostimulator are provided in the Supplementary Appendix and Supplementary figure 5. The neurostimulator was connected to two subdural electrode strips (Subdural leads, Medtronic; electrode diameter 4 mm, interelectrode distance 1 cm, 4 electrodes per lead) approved for both recording and stimulation. Positioning on target location was guided by neuronavigation. During the clinical implantation surgery, four burr holes had been made in the trepanation margin approximating a rectangle acting as a neuronavigation reference. Location of the sensing and stimulation electrode strips were marked on the cortex with a marking pen (type 1041, SandelMedical) and the neuronavigation wand. Both subdural strips were fixated to the cortex with Tisseel and each lead was sutured to the dura. During closure of the dura, the strips were fixated to the dura with sutures after verification of the correct location with the neuronavigation wand. Extension leads were connected to the electrode strip leads, tunneled subcutaneously and connected to the neurostimulator that was placed subcutaneously beneath the clavicle. The patients were discharged from the hospital 2-4 days after implantation of the neurostimulator.

### Data collection phase

During three months after implantation, we asked the patients to initiate a recording of seizure data in time domain format when they experienced a seizure (see Supplementary Appendix and Supplementary figure 5) and to simultaneously keep a seizure diary. During each research visit (one visit per two weeks), the ECoG data was exported from the neurostimulator. This data was analyzed to select frequency bands that changed significantly when pre-ictal ECoG signals changed towards the ictal state. The device was then programmed to record power domain data simultaneously with time domain data to affirm that a detectable change in power was observed during seizure onset. From this power domain data, a linear discriminant algorithm (LDA) was constructed to distinguish seizure onset activity from interictal activity. This LDA was then tested and tuned until a sensitivity of > 50% and a false detection rate of < 20/hour was reached. We then continued to the next phase in which cortical stimulation is initiated when a seizure is detected. Details of the calculation of the LDA are provided in the Supplementary Appendix and Supplementary figure 6.

### Closed-loop cortical stimulation phase

After the data collection phase, the patient was asked to continue recording seizures to verify the performance of the LDA detector and to keep a seizure diary. The patient visited the hospital once per month. For nine months, we optimized stimulation parameters to reduce seizure frequency. We compared the seizure frequency in month 11-12 with the seizure frequency during the data collection phase. Statistical analysis was performed with the Mann-Whitney U test (p<0.05).

### Quality of life, sensorimotor function and participation in society

One day before the start of the clinical monitoring period, and one year after inclusion in this study, the patient completed two questionnaires regarding the quality of life (aQoL-8D) and participation in society (USER-test). We also tested motor hand function with the Action Reach Arm Test (ARAT), the nine-hole peg test, and performed physical examination.

## Results

### Subjects

We included seven subjects in this study between November 2019 and November 2020 (see Table 1 for subject characteristics, and Figure 1 for a timeline). In two subjects (REC2Stim02 and REC2Stim04), the SOZ turned out to be located outside essential eloquent cortex, so these subjects underwent epilepsy surgery and were excluded from this study. The other five subjects (REC2Stim01, REC2Stim03, REC2Stim05, REC2Stim06, REC2Stim07) proceeded with implantation of the neurostimulator. Additional details regarding the exact implantation location of the subdural electrodes is provided in the Supplementary Appendix and Supplementary figure 7.

**Table 1:** subject characteristics. REC2Stim02 and REC2Stim04 were excluded from this study, since the SOZ was located outside the primary sensorimotor cortex and epilepsy surgery was performed. F = female, M = male, R = right, L = left.

| Subject | Sex | Affected side | Involved extremity | Resection/neurostimulator |
| --- | --- | --- | --- | --- |
| REC2Stim01 | F | R | Hand | Neurostimulator |
| REC2Stim02 | F | R | Hand | Resection |
| REC2Stim03 | M | R | Leg | Neurostimulator |
| REC2Stim04 | F | L | Mouth | Resection |
| REC2Stim05 | M | L | Leg | Neurostimulator |
| REC2Stim06 | F | R | Leg | Neurostimulator |
| REC2Stim07 | M | L | Hand | Neurostimulator |

**Figure 1:**
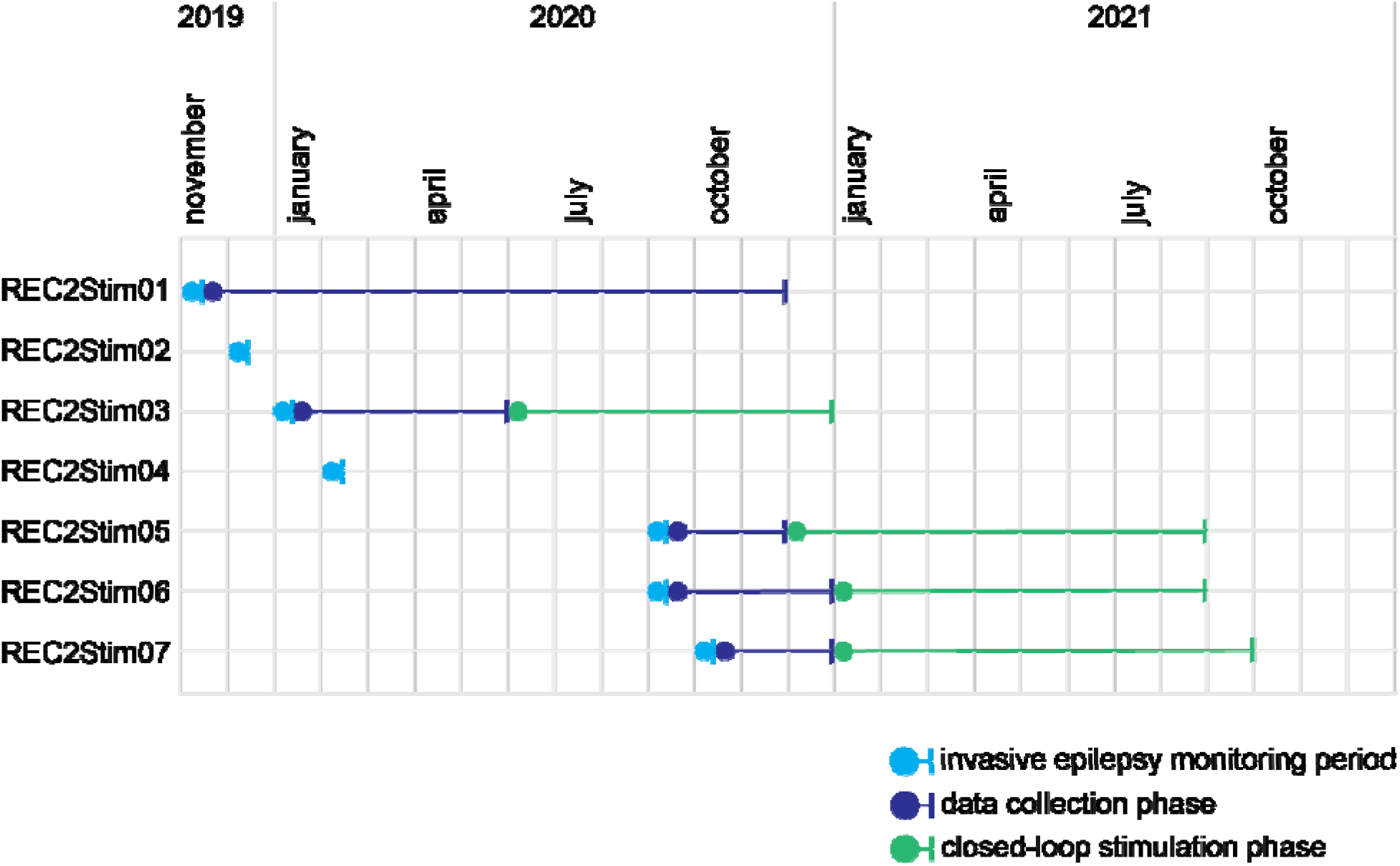
Timeline of study participation for each subject. The first subject was included in November 2019. The last subject was included in October 2020. Two subjects (REC2Stim02 and REC2Stim04) were excluded from this study during the invasive epilepsy monitoring period (light blue), since epilepsy surgery was performed. Five subjects underwent invasive epilepsy monitoring (light blue), implantation of the neurostimulator, after which the data collection phase (dark blue) and the closed-loop stimulation phase (green) followed. Study participation of the last subject ended in October 2021.

### Subjects who underwent epilepsy surgery

In REC2Stim02, we delineated the SOZ outside the primary sensorimotor hand area. The SOZ was located in the frontal cortex with fast spreading into the primary sensorimotor cortex. She underwent cortectomy in the posterior frontal lobe, anterior of the precentral sulcus. Pathology of the resected tissue showed a Focal Cortical Dysplasia (FCD) 2B. A year after surgery, she remains seizure free and will start tapering off anti-seizure medication.

In REC2Stim04, an abnormally large sensorimotor mouth representation was found in the ventral precentral gyrus. The large FCD 2B, already visible on MRI, was localized in this area and deemed resectable. One year after surgery, she still had seizures and she underwent a second resection for residual FCD on MRI. Currently, she is not completely seizure free, but the final result is entirely satisfactory for her; she went from seven seizures per night to one very short seizure a week, lasting a few seconds, on awakening.

### Data collection phase

Following the implantation of the neurostimulator, REC2Stim01 ceased to have her regular seizures, though she reported some erratic twitches in her hand. This was insufficient to optimize a seizure detection algorithm and apply closed-loop cortical stimulation. The other four subjects went on to participate in the data collection phase and stimulation phase.

During the data collection phase, we recorded on average 281 (range: 115-743) seizures per subject in the remaining four subjects. The performance of the LDA to detect seizures had a sensitivity of 70-95% and a false detection rate of 1-5/hour. Additional details regarding the performance are provided in Supplementary Appendix and Supplementary figure 8.

### Seizure frequency

In the last two months of the stimulation phase, the seizure frequency was reduced by 70%, 77%, 26% and 44% in REC2Stim03, REC2Stim05, REC2Stim06 and REC2Stim07 respectively, as compared to the seizure frequency during the data collection phase (see Figure 2). This reduction was significant in two subjects (REC2Stim03 and REC2Stim05, p<0.001).

**Figure 2:**
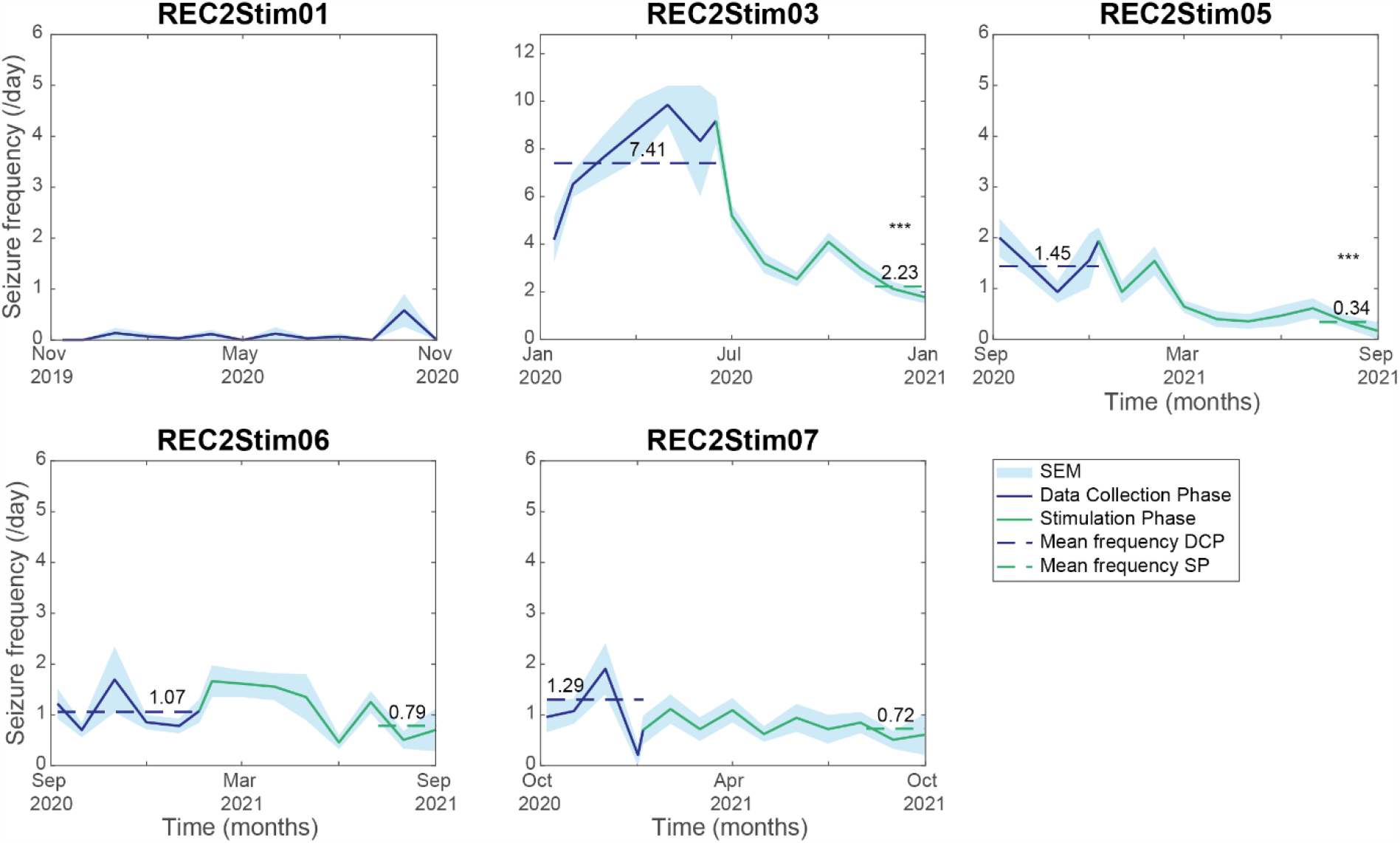
The mean seizure frequency during the data collection phase (DCP, dark blue), during the closed-loop cortical stimulation phase (SP, green) and the standard error of the mean (SEM, light blue) are displayed. The mean seizure frequency during the last two months of the SP (dotted green line) was significantly lower in subjects REC2Stim03 and REC2Stim05 compared to the mean seizure frequency during the DCP (dotted dark blue line). Note that the y-scale of REC2Stim03 has higher limits than the y-scales of the other patients. ***: p<0.001

### Quality of life, participation in society and sensorimotor function

We did not find a clear difference between quality of life before implantation of the neurostimulator and a year after study participation (see Figure 3A). In REC2Stim01 and REC2Stim06, the self-reported ability to participate in society was increased a year after study participation (see Figure 3B). In the other subjects, we did not find a clear difference. Regarding functioning of the hand (see Figure 3C&D), we did not find a clear difference. Although we did not find any differences in quality of life or ability to participate in society, we observed some individual improvements. In REC2Stim06, one anti-seizure medication was stopped because of side-effects. This did not lead to an increase of seizures. Both REC2Stim06 and REC2Stim07 had a history of yearly admissions to the hospital because of a cluster of uncontrollable seizures. This did not occur during participation in this early feasibility study. All participants expressed that they would like to continue with the closed-loop cortical network stimulation treatment after the end of study participation.

**Figure 3:**
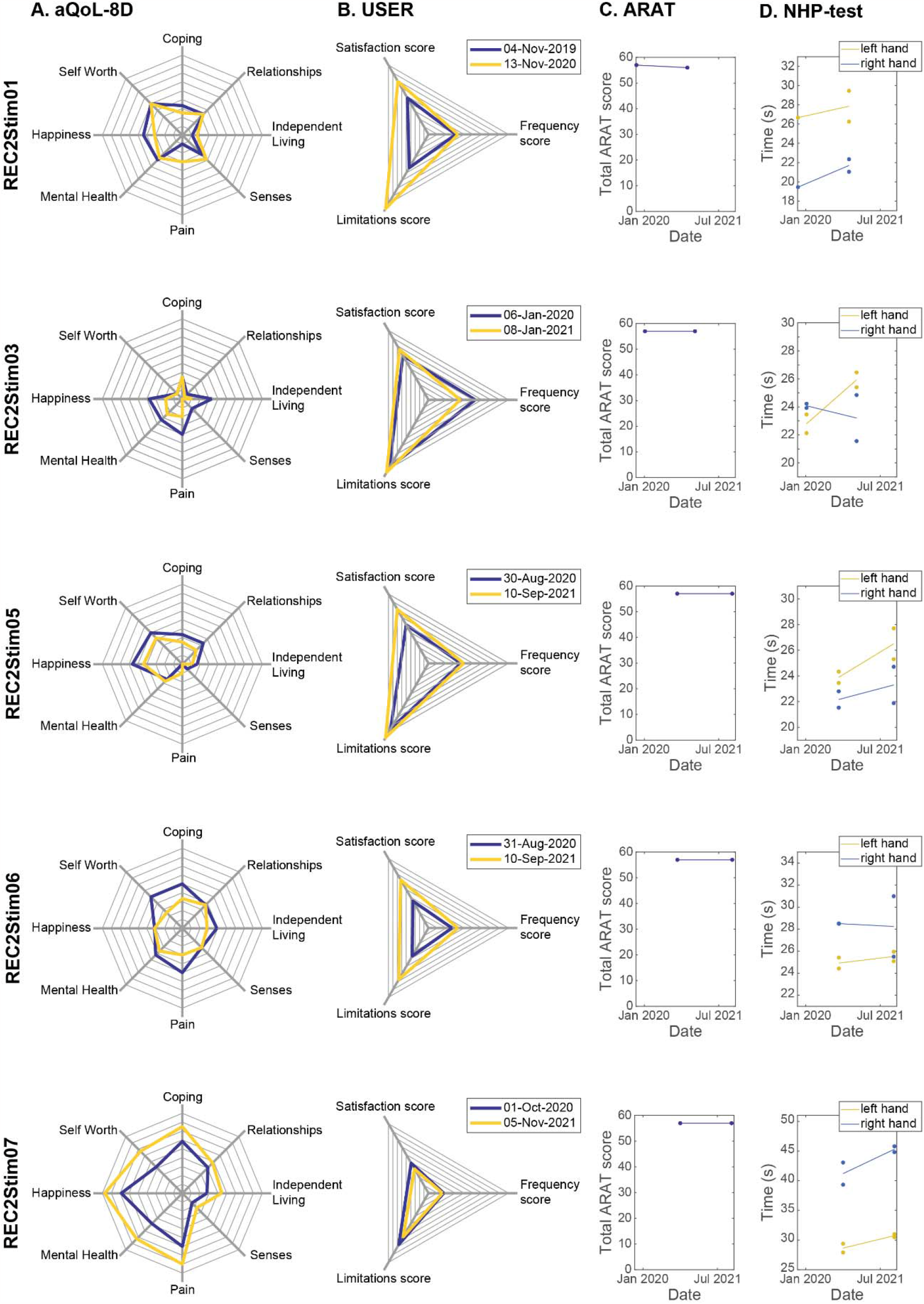
For five subjects, the quality of life, measured with the aQoL-8D (A) and the level of participation in society, measured with the USER test (B) are displayed prior to implantation of the neurostimulator (dark blue) and a year after study participation (yellow). Motor hand function, measured with the ARAT (C) and the nine-hole peg test (D), are displayed prior to implantation of the neurostimulator and a year after study participation.

### Complications

REC2Stim01 reported a headache a week after implantation of the neurostimulator. This resolved in three weeks. She also reported a tingling sensation in the left side of her tongue when she was tired. Two months later, this was resolved without intervention.

REC2Stim03 had an increase of seizure frequency (normally 1 tonic-clonic seizure per 2 weeks, and after implantation 1-4 tonic-clonic seizures every night) during a week after implantation of the neurostimulator. Clobazam was given for one week, and the seizure frequency decreased to baseline afterwards.

When we started the closed-loop cortical network stimulation, REC2Stim05 reported seizures during the day, while he was only familiar with nocturnal seizures. We resolved this by switching off the neurostimulator during the day. No other adverse events were reported.

### Other relevant findings after one year follow-up

After one year of follow-up, some relevant findings and technical complications occurred. These findings are discussed in detail in the Supplementary Appendix.

## Discussion

We implanted a neurostimulator in five subjects with refractory epilepsy arising from the primary sensorimotor cortex. One subject became seizure-free after only implanting the neurostimulator without applying any electrical stimulation. She had a presurgical high burden of regular seizures and had shown 80 seizures in six days of invasive monitoring just before. This surprising effect might be mediated by the expectation of the clinical benefit to be obtained, as was described in a study investigating placebo effect during deep brain stimulation treatment in patients with Parkinson’s disease ^15^. In the other four subjects, we were able to detect seizures with a sensitivity of at least 70% and a false detection rate of <5 /hour. Two subjects responded to closed-loop cortical network stimulation with a mean seizure frequency reduction of 73%. The two other subjects had a mean reduction of 35%. In another study applying responsive neurostimulation, the median seizure frequency reduction was 44% after one year ^16^. This reduction increased to 53% after two years and 75% after nine years ^4^. This suggests that underlying network excitability changes due to the applied neurostimulation and efficacy increases over the years. In the next few years, we will be able to evaluate whether this effect is also present in the subjects that participated in our study.

In this clinical early feasibility study, we included seven subjects suspected of focal epilepsy arising from the primary sensorimotor cortex with low odds of proceeding towards epilepsy surgery. During presurgical evaluation with subdural electrode grids covering the presumed epileptogenic regions, we concluded that epilepsy surgery was possible in two subjects (almost 30%). Prior to this study, the odds for epilepsy surgery was estimated at <10%, and clinical invasive monitoring would likely not have been done. Our study shows that epilepsy surgery might be feasible in more patients with a suspected focus in the primary sensorimotor cortex. In a study of the subjective effects of responsive neurostimulation ^17^, quality of life improved in 44% of the patients. Interestingly, these findings were not explained by changes in seizure frequency or anti-seizure medication. We did not see clear differences in quality of life pre-implantation and a year after. Our study was executed between November 2019 and November 2021. Around the same time, covid-19 impacted our daily lives. This might have influenced the quality of life ratings and ability to participate in society for our subjects. Furthermore, improvement in quality of life continues to be observed throughout a follow-up duration longer than one year ^18^.

We did not find any differences with physical examination one year after study participation compared to prior to implantation of the neurostimulator. We did not observe any differences in motor hand function due to stimulation. When setting up this trial, we expected the majority of eligible patients to have involvement of the sensorimotor hand region. In the end, three of our subjects had seizures arising from motor leg/foot area. One of the two implanted subjects with seizures arising from the motor hand area did not receive stimulation due to the absence of seizures since implantation (REC2Stim01), leaving only one subject to evaluate.

In this study, we applied closed-loop stimulation. In a large trial on responsive neurostimulation ^16^ with similar results on seizure frequency reduction, electrical stimulation was applied upon seizure detection with a burst duration of 100 ms and a total stimulation duration of 5.9 min/day, leading to a stimulation every 30 s. This suggests that not only ictal activity, but also interictal activity was detected and responded to with electrical stimulation. The question remains what is better: closed-loop stimulation, stimulation applied on both interictal and ictal activity or stimulation applied in an open-loop cycle. Some studies demonstrated reduced spike-wave-discharges in a genetic absence model in rats^19^ or suppression of seizure-like activity in hippocampal brain slices ^20^ with closed-loop stimulation that were not observed with open-loop stimulation, while high efficacy with open-loop stimulation has been demonstrated in case studies as well ^21,22^. One of the advantages of closed-loop stimulation is the minimization of side effects related to stimulation when there are no seizures ^23^. Furthermore, closed-loop stimulation minimizes power consumption and delivers a lower total daily dose of current, which both benefits battery life of the neurostimulator ^23^.

In clinical practice, treatment efficacy is commonly evaluated based on seizure diaries reported by the patient. In this study, we also relied on these self-reported seizure diaries. The seizure frequency derived from these self-reports is usually inaccurate, and does not include subclinical events ^23^, which is a general concern when evaluating treatment efficacy. Additionally to the seizure diary, subjects used a Patient Programmer (see Supplementary Appendix and Supplementary figure 5 for properties of the neurostimulator) that logged ictal events in the neurostimulator. This might produce a more reliable diary than a traditional self-reported one ^24^. During intracranial monitoring, we performed some extra stimulation trials in which we applied several stimulation frequencies and analyzed the effect in the SOZ in order to determine the stimulation site with best modulating effect on seizure activity. When starting the closed-loop cortical network stimulation phase, we selected our first stimulation frequency based on the responses to stimulation during the extra stimulation trials. We hoped that spectral changes in interictal activity due to stimulation would be a predictor for long-term neuromodulatory effects. However, in this small set of subjects, we were not able to find a clear relationship between the responses to stimulation in the extra stimulation trials during the intracranial monitoring and long-term effect on seizure frequency. This means that more research is needed to find predictors for effective stimulus parameters in the individual subject. This could minimize the long trajectory of trial-and-error with stimulus parameters that is now often clinical practice for patients with epilepsy receiving neuromodulation therapy.

In this clinical early feasibility study, we have demonstrated that closed-loop cortical network stimulation in an area of healthy tissue connected to the SOZ led to a mean seizure frequency reduction of 54%. In the following years, we will continue applying stimulation with different stimulation paradigms to improve the treatment with optimized seizure frequency reduction.

## Data Availability

All data produced in the present study are available upon reasonable request to the authors

ARAT: action reach arm test
CCEP: cortico-cortical evoked potentials
DCP: data collection phase
ECoG: electrocorticography
FCD: focal cortical dysplasia
LDA: linear discriminant algorithm
M1: primary motor cortex
REC2Stim: rational extra-eloquent closed-loop cortical stimulation
SEM: standard error of the mean
SP: stimulation phase
SPES: single pulse electrical stimulation
SOZ: seizure onset zone
USER test: Utrecht scale for evaluation of rehabilitation-participation

## Acknowledgement

Research reported in this publication was supported by EpilepsieNL under Award Number NEF17-07 (DvB) and NEF19-12 (DvB, SB). Furthermore, this research would not have been possible without the help of technicians, nurse specialists, nurses and other members of the clinical epilepsy team. We would like to thank the Data Safety Monitoring Board (chair: prof. dr. K. Vonck, members: drs. J. Ardesch, dr. L. Wagner, dr. R. Schuurman) for their critical and inspiring comments while monitoring the safety of this study. We thank Medtronic for donating devices and dr. Gaetano Leogrande for technical support. Finally, we would like to thank the patients who participated in this study.

