## Supplemental Material for "Closed-loop Cortical Network Stimulation as treatment for refractory epilepsy originating from the primary motor cortex"

***Invasive epilepsy monitoring period – Electrical stimulation mapping***

We applied electrical stimulation mapping (50 Hz, 1-6 mA, 1025 µs, biphasic, for 1-5 s) in each neighboring electrode pair. When the patient would experience symptoms like twitches, sensations or an epileptic aura, this electrode pair would be noted as involved with this specific function or with evoking seizures. After applying stimuli to each pair, we reconstructed a map that showed which areas were involved in which function and/or epileptic aura. This map facilitated delineation of regions that were suitable or unsuitable for resection, optimizing the chances of seizure freedom while minimizing the risk of neurological deficits (see Supplementary figure 1).

***Invasive epilepsy monitoring period – Single Pulse Electrical Stimulation***

We applied Single Pulse Electrical Stimulation (SPES; ten monophasic, bipolar stimuli of 0.2 Hz, 4-8 mA, 1 ms) to each adjacent electrode pair (SD LTM STIM Cortical Stimulator, Micromed, Treviso, Italy). For each stimulus pair, we selected epochs of the data time-locked to the stimulus artefact in a time window of 2 s before until 2 s after each stimulation for each response electrode.

First, we analyzed the underlying cortico-cortical network by evaluating the Cortico-Cortical Evoked Potentials (CCEPs, see Supplementary figure 1A). We averaged the epochs during ten trials per stimulus pair. In the averaged signal, a CCEP was detected when the signal exceeded 2.6 * standard deviation, which was calculated in the time window of 1 – 0.1 s prior to the stimulus artefact. All detected CCEPs were visually checked (DvB) to reduce false-positive detections.

Secondly, we analyzed which Single Pulse Stimuli modulated the SOZ by evaluating transient power reduction post-stimulation. After making epochs as described earlier, we calculated event-related spectral perturbations (ERSP ^1^) with a 3-cycle wavelet with a Hanning-tapered window in which the number of cycles increases with 20% between the frequency range of 10-250 Hz. Bootstrapping was applied to display only significant differences (p < 0.05) in power post-stimulus compared to pre-stimulus (see Supplementary figure 1B).

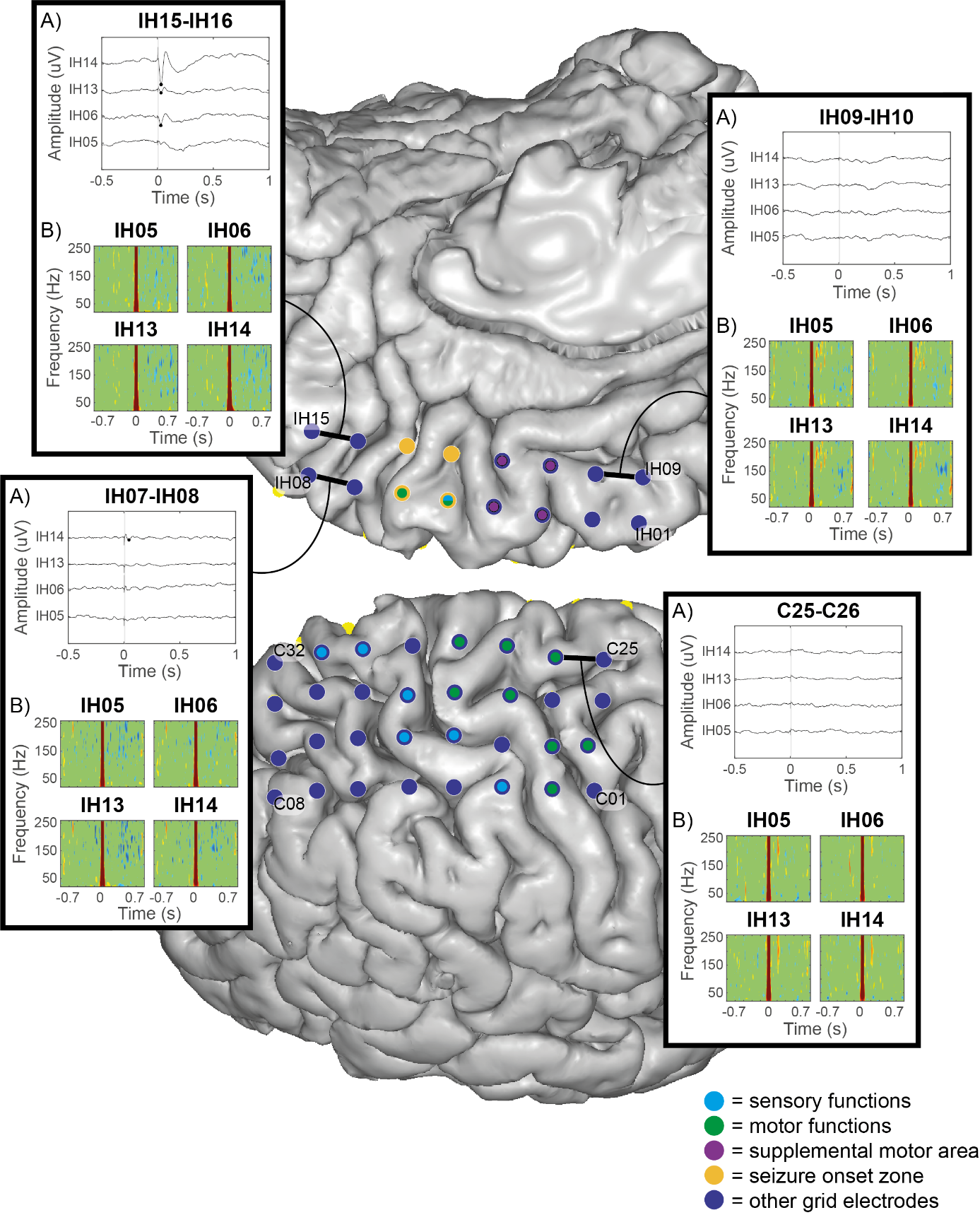

**Supplementary figure 1: Intracranial grid implantation of REC2Stim03. With electrical stimulation mapping, we delineated sensory (light blue), motor functions (green) or supplemental motor area (purple). The seizure onset zone (SOZ, yellow) was delineated based on seizures that occurred during the invasive epilepsy monitoring period. With SPES, we reconstructed connections towards the SOZ. In the upper left box, stimulation in electrodes IH15-IH16 showed a CCEP (A) in response electrodes IH6, IH13, IH14. The time-frequency plots (B) display changes in power in electrodes IH05, IH06, IH13, IH14 after stimulating IH15-16 (upper left box), IH07-IH08 (lower left), IH09-IH10 (upper right) and C25-C26 (lower right). We did not observe any CCEPs or transient power suppression in electrodes IH05, IH06, IH13, IH14 when stimulating electrode pair C25-C25. We observed either CCEPs or transient power suppression in IH05, IH06, IH13, IH14 when stimulating any of the other three electrode pairs.**

***Selection of seizure detection site***

Seizures were visually annotated by the responsible neurologist (FL) and the clinical neurophysiology team. We selected epochs with a time window of 30 s pre-seizure to 30 s after seizure onset of all seizures that occurred spontaneously during the invasive epilepsy monitoring period. We applied a notch filter (Butterworth, 3^rd^ order, 47-53 Hz and 97-103 Hz) to remove 50 Hz and 100 Hz line noise. We applied a Gabor wavelet convolution and calculated the power in the frequency bands 4-7 Hz, 8-14 Hz, 15-25 Hz, 26-40 Hz and 65-95 Hz in a time window of 15 s pre-seizure and 5 s during seizure onset. We averaged the power in each frequency band over all samples pre-seizure and during seizure onset and tested statistical significance between the power pre-seizure and during seizure onset in each frequency band in each electrode with a Wilcoxon signed rank test (p < 0.05). We applied FDR correction to correct for multiple testing (see Supplementary figure 2). The electrodes that showed the largest difference between interictal and ictal power spectra were selected as the sensing site for seizure detection.

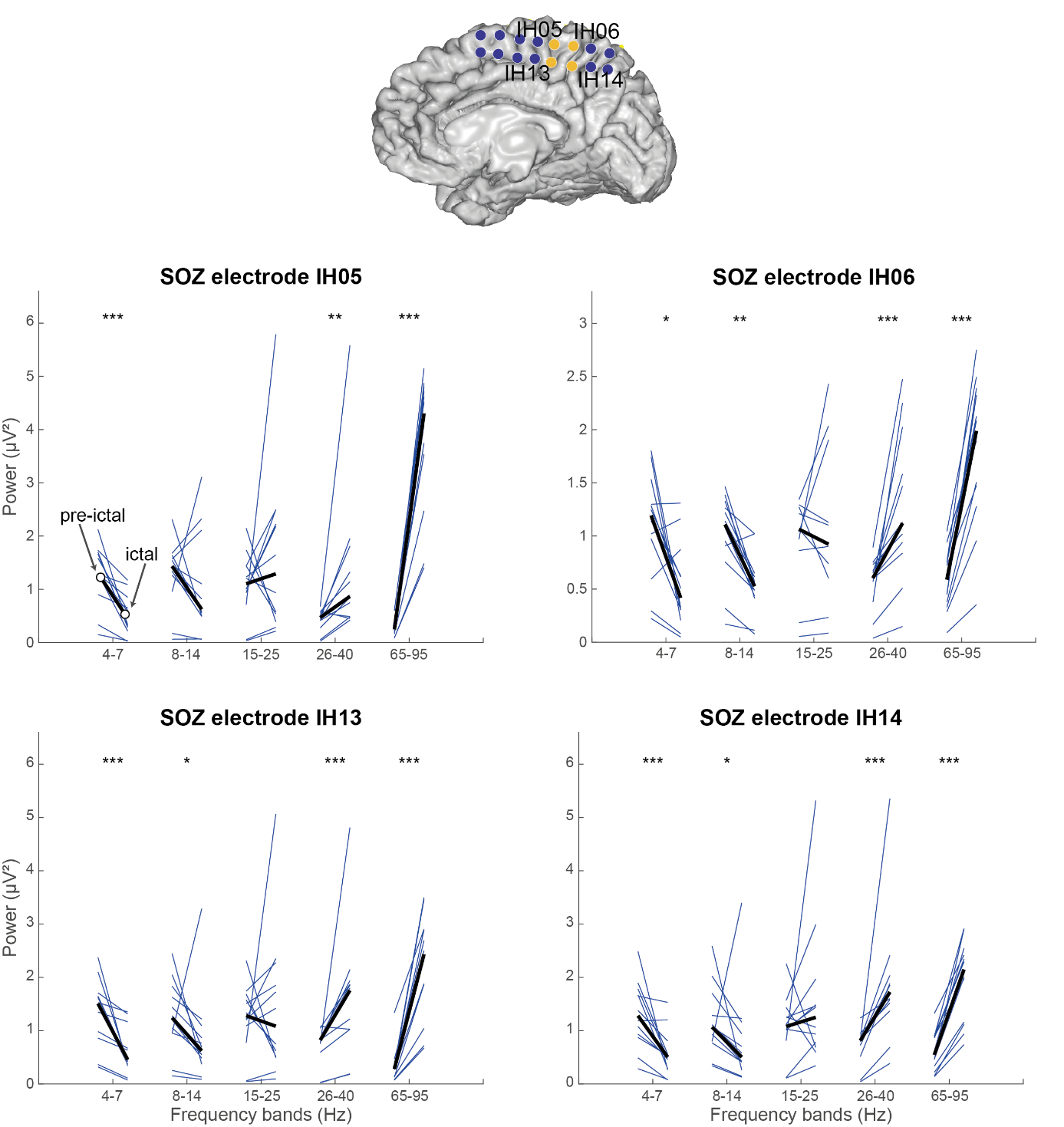

**Supplementary figure 2: Selection of electrodes for seizure detection in REC2Stim03. In each frequency band, the power in pre-ictal (left) and ictal (right) epochs are displayed. In all electrodes defined as covering SOZ, we observe that there is a power increase in 26-40 Hz and 65-95 Hz during ictal activity. Power is decreased in 4-7 Hz frequency band during ictal activity. In all electrodes except IH05, a decrease in power in 8-14 Hz frequency band was also observed during ictal activity. *: p<0.05, **: p<0.01, ***: p<0.001 (FDR corrected).**

***Selection of electrodes for therapeutic stimulation***

We selected three potential candidates for extra stimulation trials based on whether the potential stimulus sites evoked a CCEP in the SOZ and whether we observed transient power suppression in the SOZ after SPES stimulation (see Supplementary figure 1 for more details).

We applied runs of ten stimuli (5 s of stimulation, 25 s rest) at the current intensity that did not evoke after-discharges (3-15 mA) during electrical stimulation mapping, and a pulse width of 120 µs at various stimulation frequencies (2, 7, 100, 130, 200 Hz). After each set of ten stimuli, we paused for ten minutes and continued with the next set of ten stimuli. In total, this protocol took five hours (see Supplementary figure 3A). Due to limited time in this invasive epilepsy monitoring period, we could only select three potential stimulation candidates.

One day prior to the implantation of the neurostimulator, we increased the current intensity in the selected stimulation site with steps of 0.5 mA to see what current intensity would lead to clinical symptoms or after-discharges (see Supplementary figure 3B).

During this epilepsy monitoring period, we had to analyze effects of stimulation trials on interictal data. Due to the limited time, it was not possible to analyze effects of these various stimulation trials on ictal data.

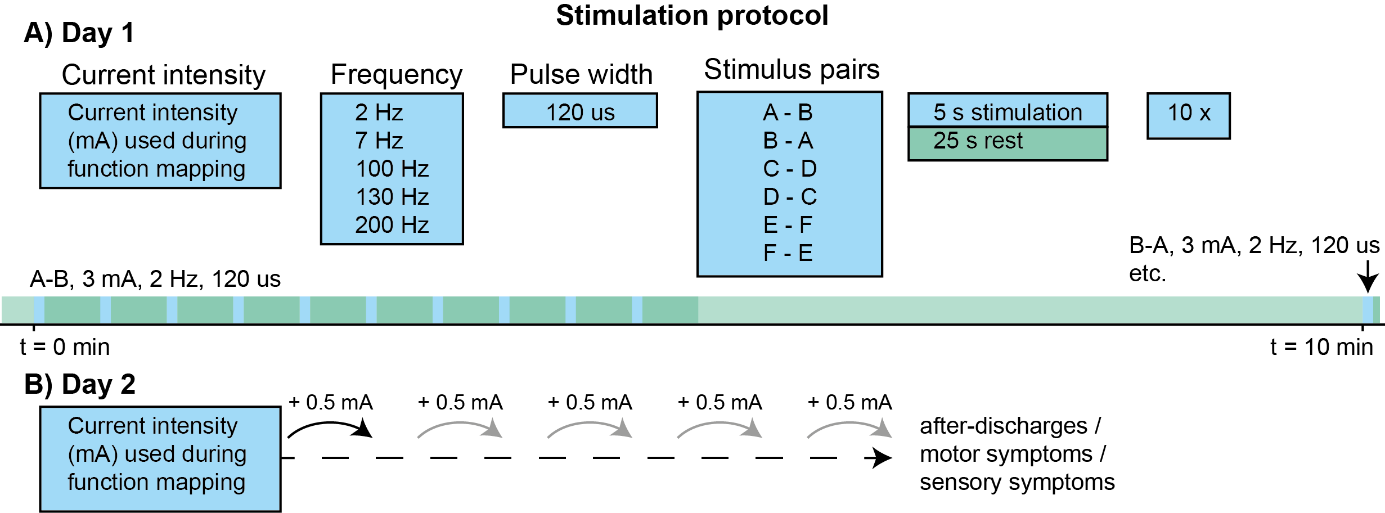

**Supplementary figure 3: The stimulation protocol was executed on two consecutive days. A) On the first day, ten stimulation trials with various stimulation frequencies were applied in three stimulation sites. B) On the second day, current intensity was increased with iterative steps of 0.5 mA to determine the potential range of stimulation without functional effects or evoking after-discharges.**

Data was recorded with a sample frequency of 2048 Hz. We re-referenced the data with a common average. The electrodes in which artefacts or 50 Hz line noise was observed, were excluded in this common average. We also applied stimulus artefact removal with the following method. We calculated the average of the signal at two sample points: 10 samples before stimulus onset and 30 samples after stimulus offset and replaced the data with this average value in a time window of stimulus onset until 20 samples after stimulus offset. The stimulus artefact would otherwise leak into the epochs that we wanted to analyze. We then applied a notch filter (Butterworth, 3^rd^ order, 47-53 Hz and 97-103 Hz) to remove 50 Hz and 100 Hz line noise.

We selected epochs of 45 s pre-stimulation and 45 s post-stimulation, applied a Gabor wavelet convolution and calculated the power in the frequency bands 4-7 Hz, 8-14 Hz, 15-25 Hz, 26-40 Hz and 65-95 Hz in a time window of 11-1 s pre-stimulation and 1-11 s post-stimulation. We averaged the power in each frequency band over all samples pre- and post-stimulation and tested statistical significance between the power pre- and post-stimulation in each frequency band for each stimulation parameter with a Wilcoxon signed rank test (p<0.05). We applied FDR correction to correct for multiple testing (see Supplementary figure 4). Based on the effect of applying stimuli in the three stimulation pair candidates, we determined which site would be most promising for long-term therapeutic stimulation.

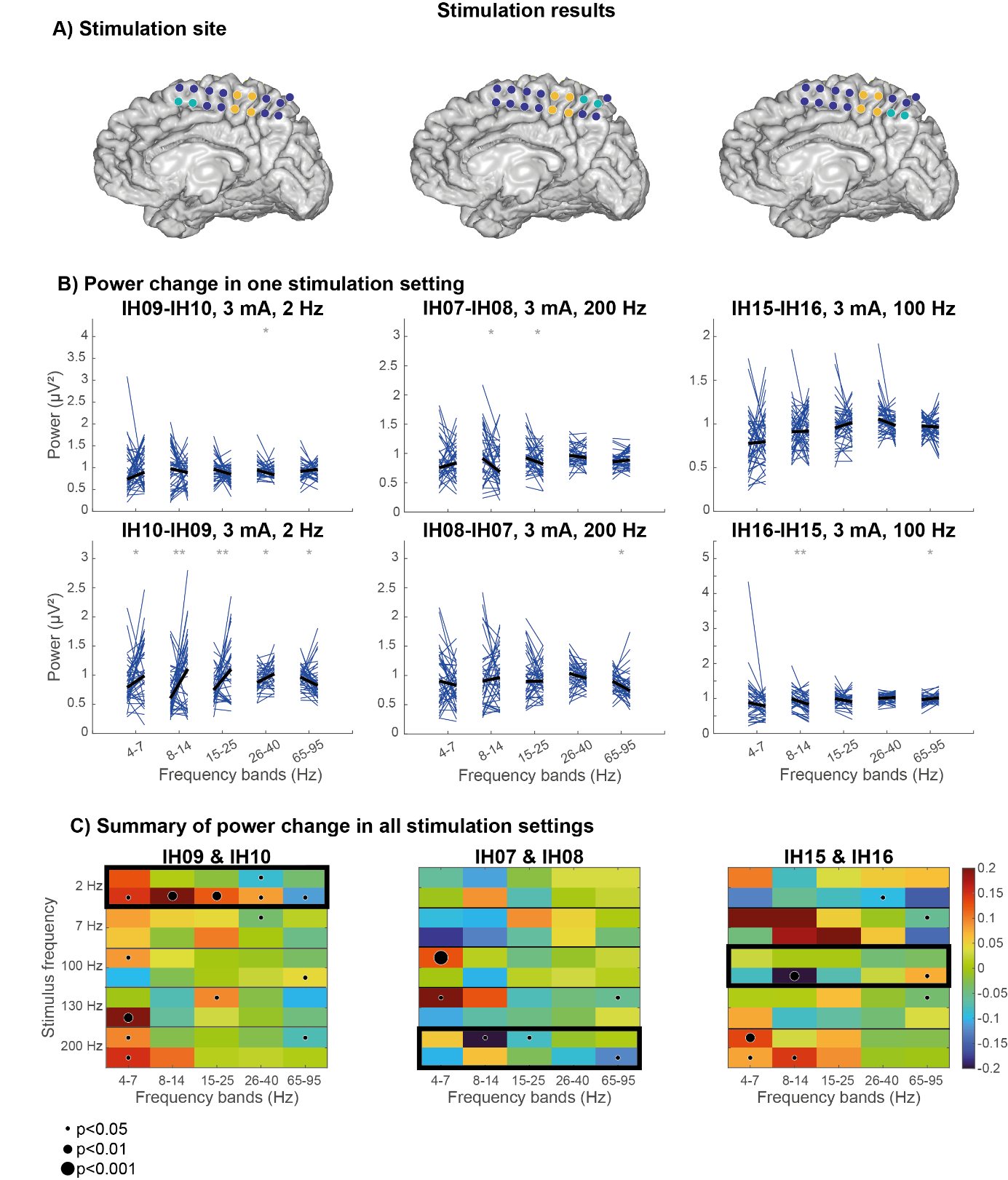

**Supplementary figure 4: Selection of electrodes for therapeutic stimulation. A) In yellow, the electrodes in the seizure onset zone were indicated. In light blue, the stimulation site is indicated. In purple, all other intracranial electrodes are indicated. B) For each stimulation site, the effect of one set of stimulation parameters is displayed for five frequency bands of the response electrodes located in the seizure onset zone. **: p<0.01, *: p<0.05 (no FDR correction, since this would remove all significant differences). C) A summary of power change in all stimulation settings. For each stimulus frequency, the two polarities of stimulation are shown (e.g. IH09-IH10 and IH10-IH09). The thick box around one stimulation frequency indicates which example was shown in B.**

***Features of the Implantable Pulse Generator (Activa***^®^ ***PC+S)***

The Activa^®^ PC+S is able to record data in three modes: 1) at a specific moment in time (e.g. every 6 hours), 2) when a patient initiates a recording by pushing a button on the Patient Programmer, 3) when a certain event is detected. In 2) and 3), data is continuously recorded in a buffer and can be stored before and after the Patient Programmer is used or an event is detected. Thus, when initiating a recording when the patient experiences a seizure, data is also stored during a few seconds prior to the use of the Patient Programmer.
During the data collection phase, we asked a patient to initiate a recording by using the Patient Programmer (see Supplementary figure 5A). We also recorded interictal data according to a time schedule. A log-file is made automatically in the Activa^®^ PC+S to track all detections, time triggered data recordings and Patient Markers. During a research visit, both the recorded data and this log-file were exported from the Activa^®^ PC+S to the Sense Programmer (see Supplementary figure 5D) via an antenna and SPTM (see Supplementary figure 5B and 5C), after which the data was transferred to an external computer. The seizure data and interictal data was used to optimize the seizure detection algorithm. Coefficients of this detection algorithm were transferred back to the Sense Programmer and implemented in the Activa^®^ PC+S.

The Clinician Programmer (see Supplementary figure 5E) was used to evaluate the battery level and impedances in the electrodes. During the cortical closed-loop stimulation period, the Clinician Programmer was used to set stimulation parameters.

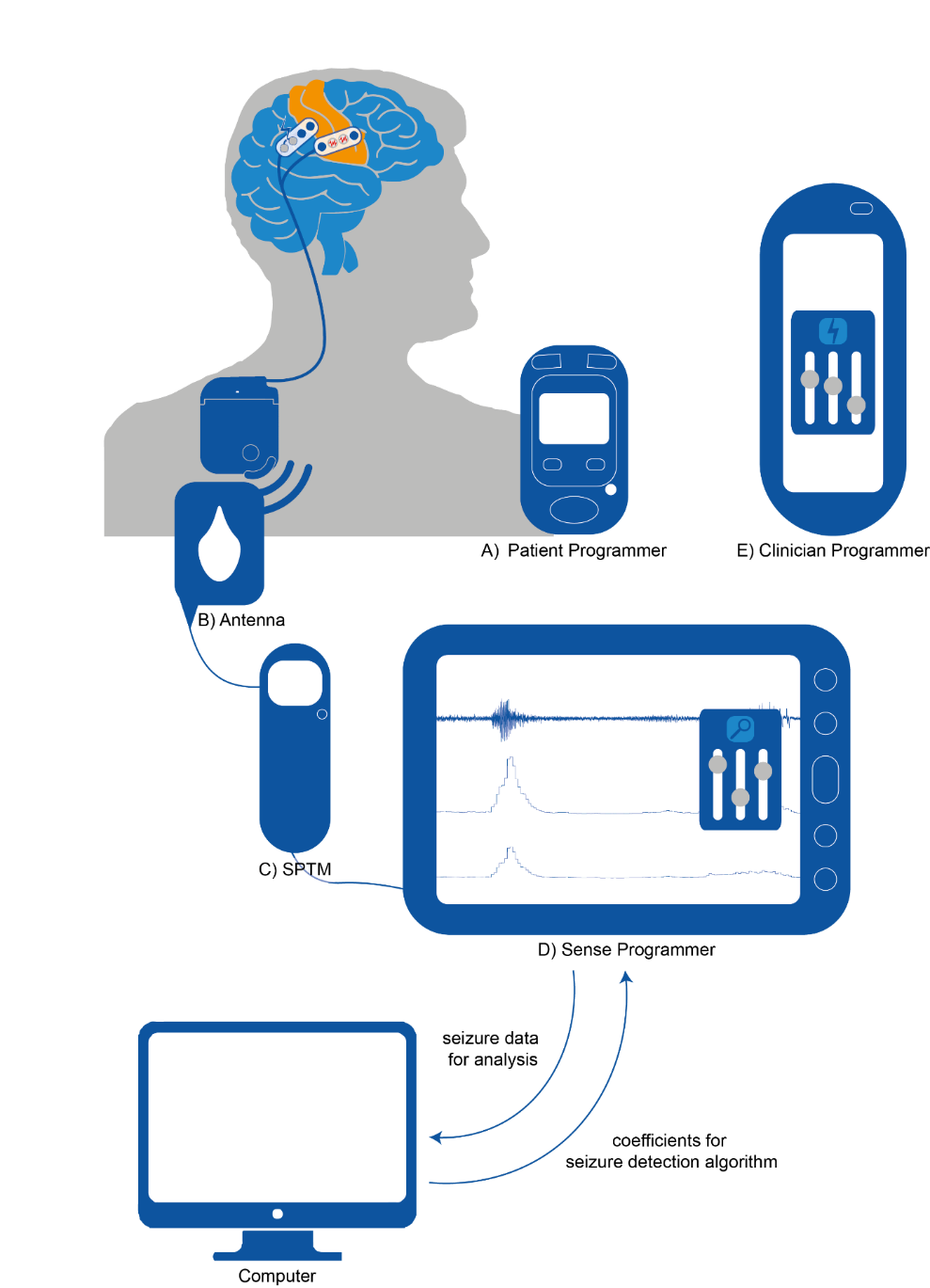

**Supplementary figure 5: components of the neurostimulation system. The subdural electrodes are placed on top of the SOZ in the primary sensorimotor cortex (orange) and on a stimulation site outside this eloquent area. These electrodes are connected to the Activa^®^ PC+S via extension leads. The patient initiates a recording by pushing a button on the Patient Programmer (A). During a visit at the outpatient clinic, the antenna (B) is placed on top of the neurostimulator and connected to the Sense Programmer (D) via the SPTM (C). The Sense Programmer is used to visualize the recordings, change sensing settings and implement the coefficients for the seizure detection algorithm. The Clinician Programmer (E) is used to check battery level and impedances of the electrodes, and set stimulus parameters.**

***Data collection phase***

Based on CT and MRI, we determined the electrode pair that covered the seizure onset zone (see Supplementary figure 6A). During the data collection phase, the subject was able to initiate the recording of seizures by pushing a button in the Patient Programmer device. With the Activa^®^ PC+S, it is possible to record one intracranial EEG signal per electrode strip in time domain. Seizures of this electrode pair were recorded as time domain data (sample frequency of 200 Hz) (see Supplementary figure 6B). Seizures were visually annotated (DvB) in Matlab R2022b. We applied a notch filter (Butterworth, 3^rd^ order, 47-53 Hz and 97-100 Hz) to remove 50 Hz and 100 Hz noise.

We selected epochs of 30 s pre-ictal to 30 s after ictal onset, applied a Gabor wavelet convolution and calculated the power spectrum in the frequencies 1-100 Hz in a time window of 10 s before seizure onset and the first 5 s of ictal onset (see Supplementary figure 6C). From this power spectrum, we determined two potential center frequencies for seizure detection. From this moment onwards, the subject initiates the recording of seizures with both time domain data and power domain data to see whether this power was increased during seizure onset (see Supplementary figure 6D). We calculated a linear discriminant algorithm (LDA) with a cost function for logistic regression (see Supplementary figure 6E). When this LDA exceeded 0 for a certain amount of samples (see Supplementary figure 6F), a detection was logged. In the stimulation phase, this detection would lead to a stimulation of a certain duration.

Hz

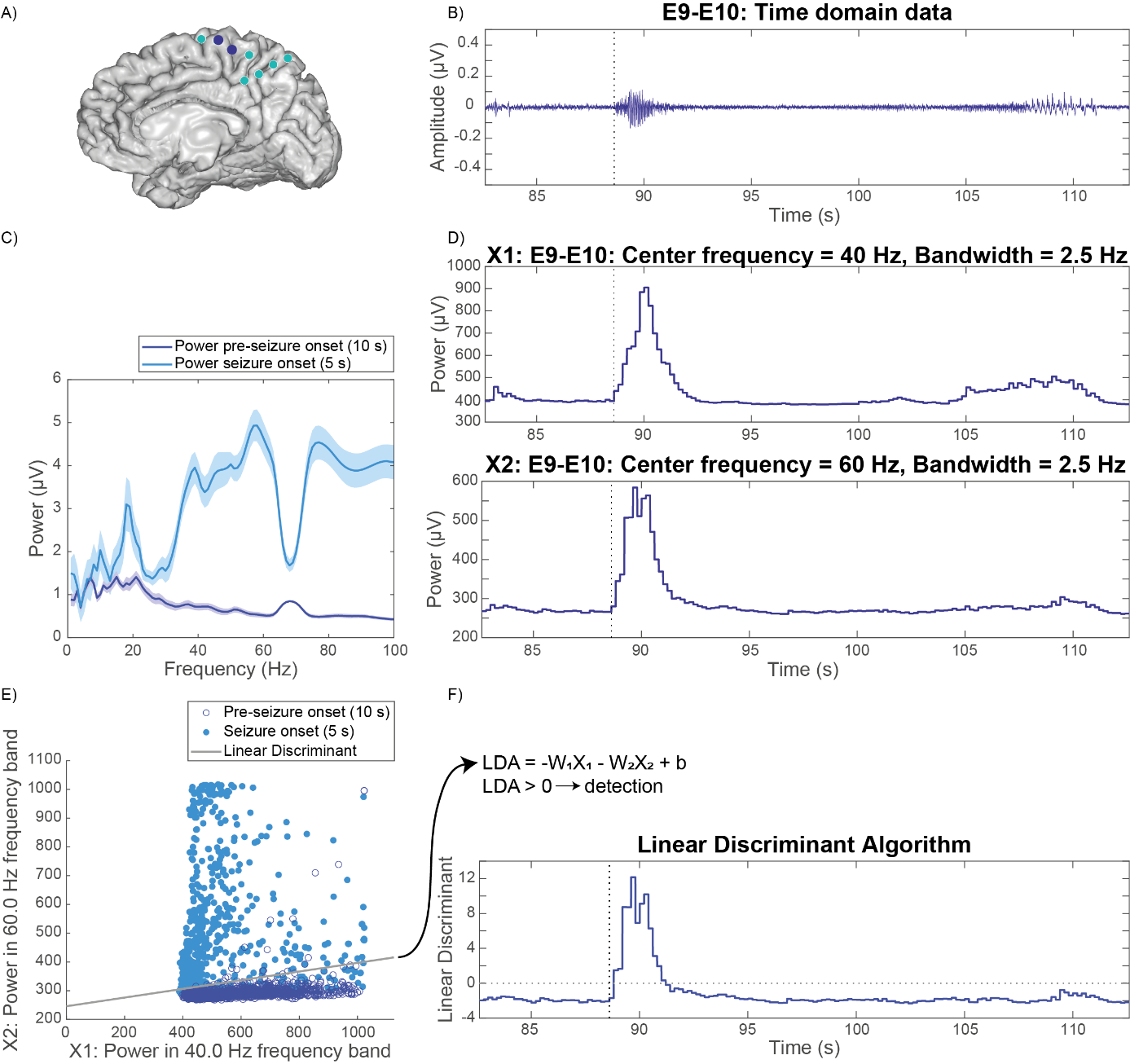

**Supplementary figure 6: Detection of seizures with a linear discriminant algorithm (LDA). A) In REC2Stim03, the subdural electrode strips (light blue) were placed interhemispherically. The purple electrodes were used for seizure detection. B) One trace of time domain data of the electrodes located on the seizure onset zone (purple in A) is displayed. The vertical dotted line at 89 s indicates the start of a seizure. C) The mean power spectrum with standard error of the mean is displayed of time domain data (displayed in B) during 10 s pre-seizure onset (dark blue) and during 5 s after seizure onset (light blue). An increase in power during seizure onset is observed from 30-100Hz, with two local peaks around 40 and 60 Hz. D) Two power domain traces (X1 and X2) are displayed of the same electrode pair as the time domain trace displayed in B). The dotted vertical line at 89 s displays seizure onset. There is a clear increase in power during seizure onset in both frequency bands. E) A scatter plot is displayed with the power in 40 and 60 Hz frequency band for each sample of the power domain traces displayed in D during 10 s pre-seizure onset (dark blue, not filled) and for each sample during 5 s after seizure onset. The cost function calculated the coefficients (W1, W2, b) of the optimal discriminant with lowest costs (light grey line). F) The linear discriminant displayed in E) is used to detect seizures. The vertical dotted line at 89 s indicates seizure onset. The LDA exceeds 0 <1 s after seizure onset and a seizure is detected.**

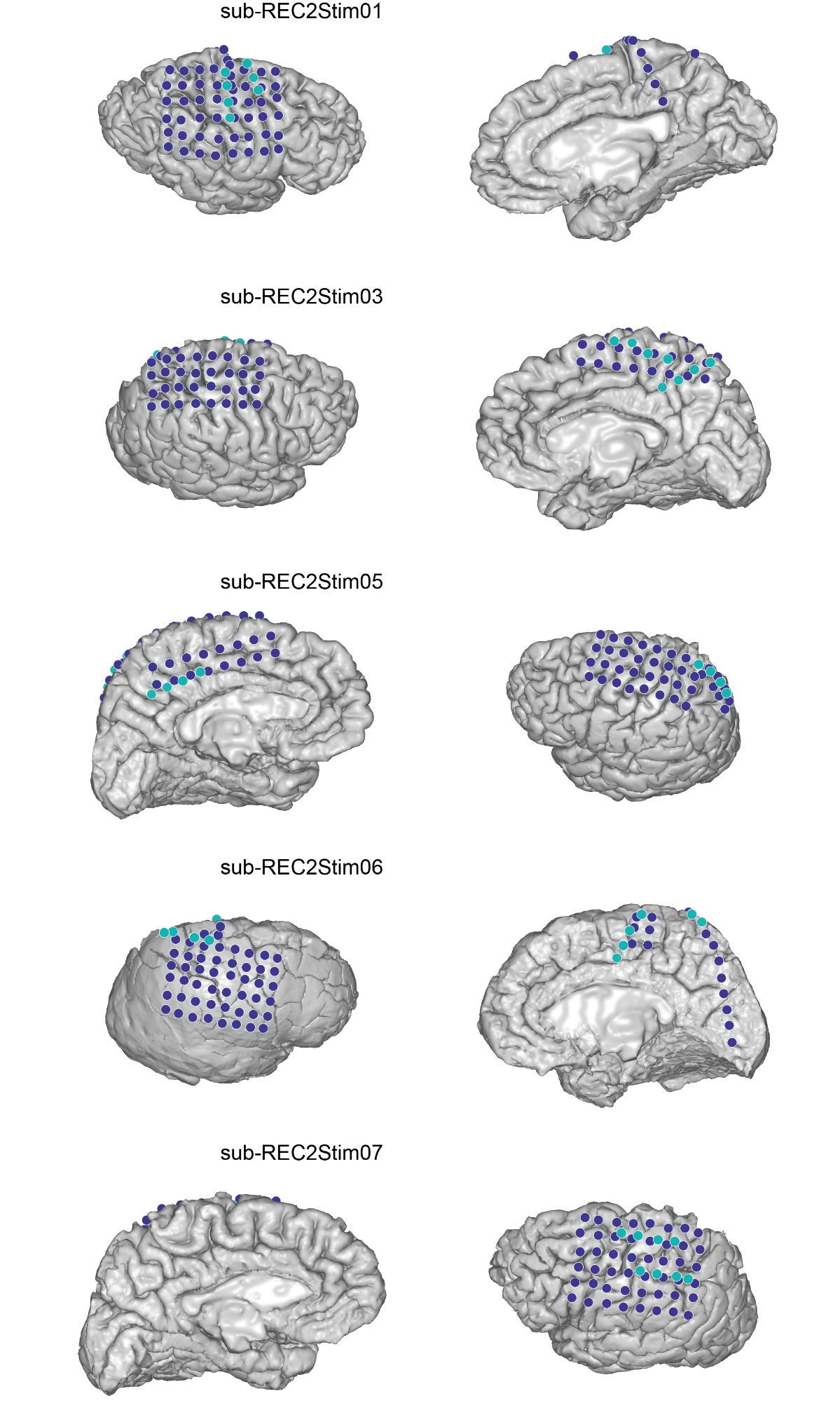

**Supplementary figure 7: For each subject with implanted neurostimulator, both the subdural grid configuration (purple) and Medtronic subdural leads (light blue) as determined with MRI and CT are displayed.**

***LDA performance***

During each visit in the data collection phase, recorded seizures were exported from the neurostimulator and analyzed to calculate sensitivity (true positive events/(true positive and false negative events)), positive predictive value (true positive events/(true positive and false positive events)) and false detection rate (false positive events/hour). If sensitivity was <50% or the false detection rate was >20 /hour, we improved the LDA (see Supplementary figure 8). The most effective LDA was used in the closed-loop cortical network stimulation phase.

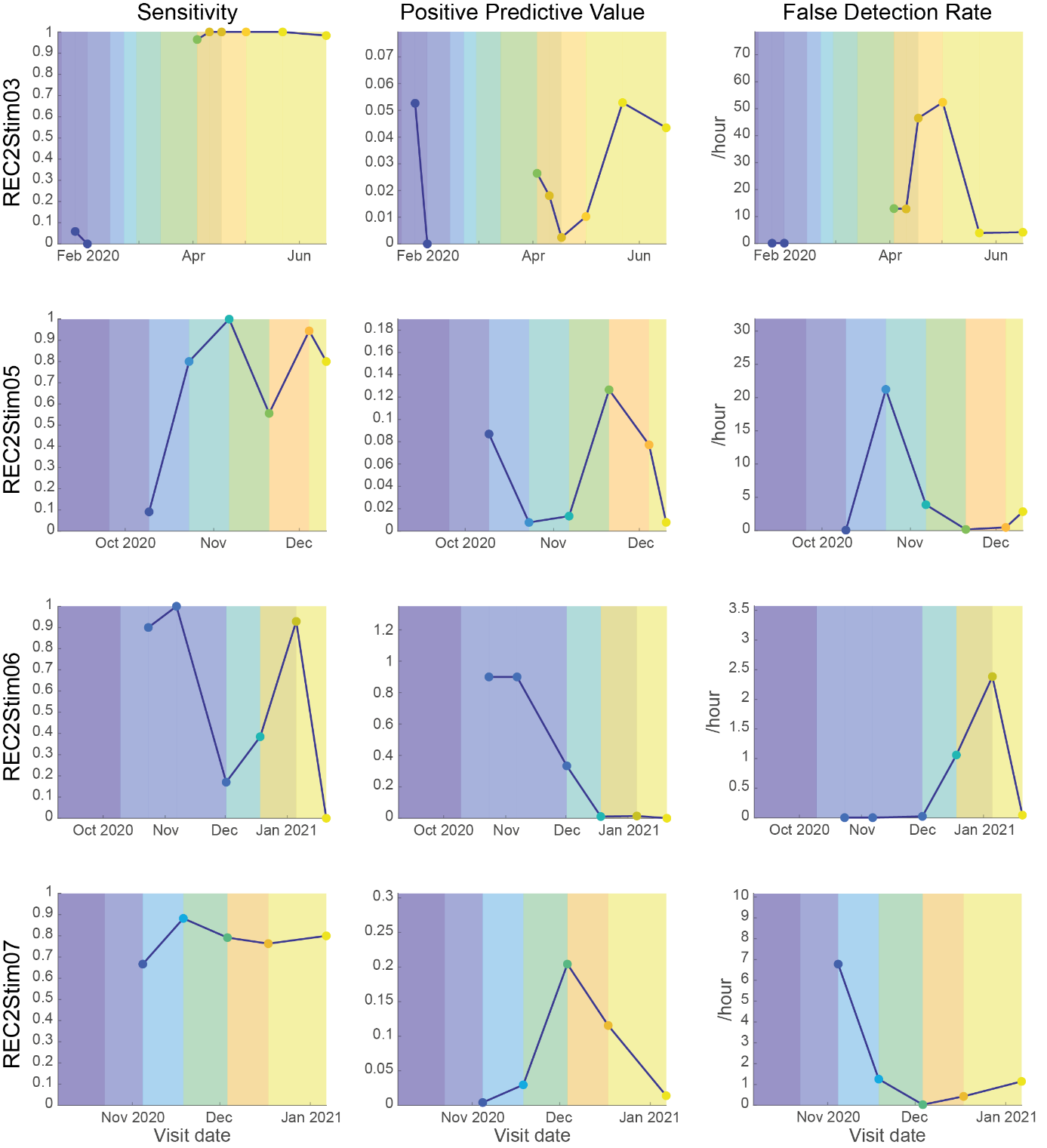

**Supplementary figure 8: Details on the performance of seizure detection during the data collection phase. Each color on the background indicates new coefficients of the LDA based on evaluation of previously recorded seizures. For REC2Stim03 eight different LDAs were evaluated before we continued to the closed-loop cortical network stimulation phase, REC2Stim06 had 4 different LDAs before we continued to the stimulation phase.**

***Other relevant findings – technical complications***

REC2Stim03 experienced a suspected broken lead which resulted in high impedances in the electrodes used for stimulation from March 2021 onwards. Stimulation therapy through these electrodes with high impedance was not effective anymore and the patient experienced an increase in seizure frequency. We changed stimulation to an adjacent electrode pair, but seizures remain present and more optimization of stimulation parameters is needed.

The Implantable Pulse Generator of both REC2Stim05 (June 2022) and REC2Stim07 (November 2021) had a software issue which led to continuous seizure detection which resulted in continuous stimulation. They were not able to turn off stimulation themselves. Both patients did not experience an increase in seizure frequency. During a research visit, we were able to turn off stimulation. When we turned on closed-loop stimulation again, this software issue disappeared and did not occur again.

***Other relevant findings – sham stimulation***

In REC2Stim03 and REC2Stim05, we initiated two weeks of sham stimulation in December 2021, because they responded well to closed-loop cortical network stimulation with seizure frequency reductions of 73%. During this period of two weeks, seizure frequency did not change. A period of two weeks might be too short to evaluate placebo effect due to an unknown duration of wash-out effect of neurostimulation ^2^.

***Battery level and impedance***

In Supplementary figure 9, the battery level and impedance of the sensing and stimulation electrode pairs are displayed. This figure indicates that closed-loop stimulation does not affect battery level significantly. We expect the battery life to have a duration of at least 4 years with similar stimulation therapy. The impedance of the sensing and stimulated electrode pairs increases in the first few months and then stabilizes.

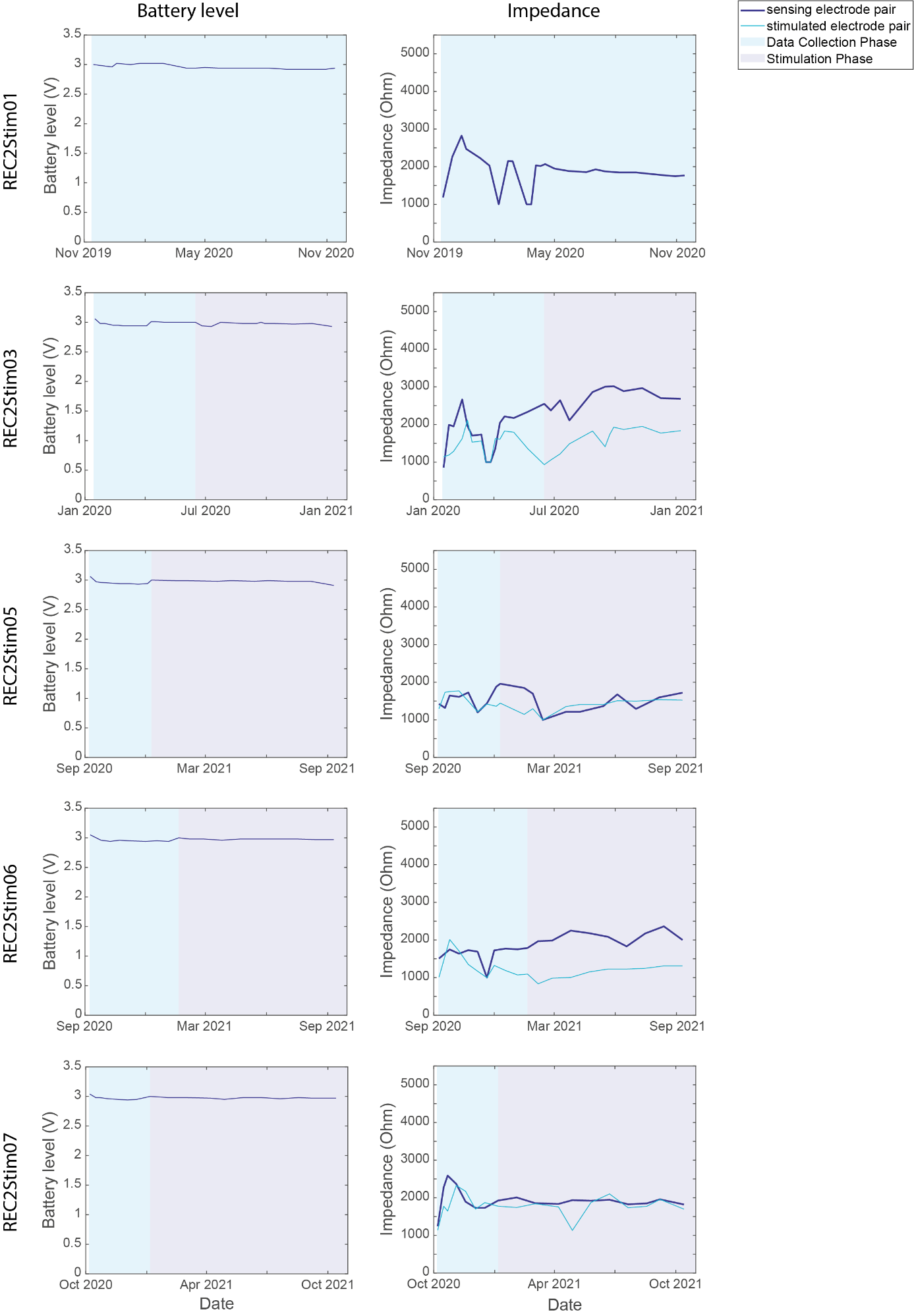
**Supplementary figure 9: Impedance and battery level of the Activa^®^ PC+S.**

***References***

1.	Delorme A, Makeig S. EEGLAB: An open source toolbox for analysis of single-trial EEG dynamics including independent component analysis. *J Neurosci Methods*. 2004;134(1):9-21. doi:10.1016/j.jneumeth.2003.10.009

2.	Valentín A, Ughratdar I, Cheserem B, et al. Epilepsia partialis continua responsive to neocortical electrical stimulation. *Epilepsia*. 2015;56(8):e104-e109. doi:10.1111/epi.13067
